# Genetic Evidence for Soluble VEGFR2 as a Protective Factor Against Macular Pucker

**DOI:** 10.1101/2024.11.29.24318222

**Authors:** Qi Gao, Jinglong Guo

## Abstract

**Background:** Identifying factors that protect against macular pucker (MP) is crucial for developing effective treatments. We hypothesize that soluble vascular endothelial growth factor receptor 2 (sVEGFR2), a decoy receptor for VEGF signaling, may provide such protection.

**Methods:** We performed a genome-wide association (GWAS) meta-analysis on three independent studies of sVEGFR2 protein quantitative trait loci (pQTL), which was used to identify novel variants and select variants strongly associated with sVEGFR2 levels. We investigated the association between sVEGFR2 levels and the risk of MP using *cis*-Mendelian Randomization (*cis*-MR) and Bayesian colocalization. We also used the UKB-PPP study of sVEGFR2 pQTL, which used a different method for measuring plasma protein, to validate the results from *cis*-MR and colocalization.

**Results:** GWAS meta-analysis identified four novel variants associated with sVEGFR2 levels, rs9411335 (*MED27*), rs6992534 (*ERI1*), rs11893824 (*LIMS2*), and rs7678559 (*PCDH7*). *Cis*-MR analysis suggested that higher levels of sVEGFR2, genetically predicted using the meta-analysis dataset, were associated with a lower risk of MP (OR 0.86, 95% CI 0.77-0.95, P = 4.6 × 10^−3^). Colocalization supported shared genetic variants in the *VEGFR2* gene region between sVEGFR2 and MP (PPH4 = 0.92). Using sVEGFR2 pQTL from UKB-PPP study, sVEGFR2’s inverse association with MP was validated by *cis*-MR (OR 0.86, 95% CI 0.78-0.96, P = 4.4 × 10^−3^) and colocalization (PPH4 = 0.95). Sensitivity analyses showed no evidence of pleiotropy or reverse causality, though some heterogeneity was found.

**Conclusions:** Our study suggested that sVEGFR2 has a protective effect against MP and could be a potential therapeutic target. Further research is necessary to elucidate its protective mechanisms and validate its translational implications.

## Background

Macular pucker (MP), also known as epiretinal membrane, is a potentially sight-threatening vitreoretinal disease. It is characterized by a sheet of abnormal, thin, fibrotic tissue that grows on the surface of the macula that is responsible for detailed central vision. The prevalence of MP ranges from 7% to 11.8%, with aging being the primary nonmodifiable risk factor. Surgical intervention, such as vitrectomy with membrane peeling, is the current standard of care for symptomatic MP cases (1–3). However, pharmacological approaches targeting MP pathogenesis are still largely lacking, likely due to an insufficient understanding of the causal factors involved (1–3).

The MP pathogenesis involves a complex interplay among various cells and acellular factors, including angiogenesis, inflammation, and fibrosis at the vitreoretinal interface (4–6). VEGFR2 is expressed in retinal pigment epithelium and endothelial cells within MP tissues, indicating its potential role in the disease’s pathogenesis (7–9). VEGFR2 signaling, which is activated through the binding of VEGF-A, promotes crucial processes in endothelial cells of blood vessels such as proliferation, migration, and survival (10). In addition, VEGFR2 signaling is also implicated in fibrosis and inflammation (11). Of note, soluble forms of VEGFR2 (sVEGFR2) can regulate VEGFR2 signaling by acting as decoy receptors for VEGF-A (12–14). In mice, sVEGFR2 has been shown to suppress corneal neovascularization and lymphangiogenesis (15). In humans, sVEGFR2 has been suggested to play an important role in macular edema through regulating angiogenesis (16, 17). Despite this, the causal relationships between sVEGFR2 and human retinal diseases such as MP remain not inferred.

Mendelian randomization (MR) is a method used to evaluate the potential causality of a modifiable exposure on an outcome by utilizing genetic variants as instrumental variables (18). Because genetic variants are randomly assorted during gamete formation and fixed at conception, MR estimates could minimize bias resulting from confounding factors or reverse causality. *Cis*-MR specifically refers to MR studies that use genetic variants from gene-restricted regions of pharmacological protein of interest, and it is typically employed for drug target validation (19). Bayesian colocalization analysis evaluates whether two distinct traits are affected by the same causal variants, making it an important complementary analysis to appraise the validity and strength of a *cis*-MR estimate (20). Meta-analysis of independent genome-wide association studies (GWAS) combines data from multiple studies into a single, large-scale dataset (21). This approach not only helps in discovering new genetic variants but also enhances the reliability and robustness of results when used in MR and colocalization studies (22). Using these advanced genetic methodologies, we investigated the relationship between plasma levels of sVEGFR2 and MP risk.

## Methods

### Data source

Information of the data sources used in this study was summarized in Supplementary table 1. The study relied on summary-level GWAS data that have been available publicly, ethical approvals were obtained in all original studies. sVEGFR2 pQTL used for meta-analysis included Interval study, with 3,301 Europeans (23); AGES study, with 5,361 Icelanders (24); and DECODE study, with 35,559 Icelanders (25). The plasma protein in these studies were measured by SomaScan platform. In addition, sVEGFR2 pQTL from the UK Biobank Pharma Proteomics Project (UKB-PPP) study (plasma protein measured by Olink and included 33,822 UK participants) was also used (26). In the original study, plasma proteins were measured with standard quality controls and adjusted for year of birth, sex, and year of sample collection. GWAS of MP (N=4,599 cases, N=413,784 controls) and other macula conditions were from the 11th release of FinnGen study, with cases being defined based on International Classification of Diseases (ICD-10). The original GWAS of eye diseases in the FinnGen study were adjusted for age, sex, and genetic principal components (27).

### Genome-wide association studies of sVEGFR2 pQTL

We performed a meta-analysis on plasma sVEGFR2 pQTL using data from three independent studies Interval, AGES, and DECODE. To account for potential heterogeneity in genetic associations, due to difference participants and/or environment, we performed the meta-analysis of random effects using METAL (version 2011-02-25) (28). This comprehensive analysis resulted in a dataset of 44,221 individuals and over 25 million variants. The base pair positions in original Interval and AGES studies were based on GRCh37, which were transformed to GRCh38 using LiftOver (version 1.28.0) (29). Finally, all the datasets used in the studies were based GRCh38. Unsing PLINK 2.00 alpha (30), independent lead genetic variants were identified by filtering associations on a genome-wide significant P-value of 5 × 10^−8^ and clumping to a R^2^ of 0.001 with 10000kb window based on the European 1000 Genome Project reference panel. The nearest protein-coding genes were identified using Open Targets, with CADD scored evaluated (31). Combined Annotation-Dependent Depletion (CADD) has been widely used to predict the deleteriousness of variants throughout the human genome (32). Novel variants were identified by excluding variants that located within 500kb of loci in original studies (Interval, AGES, and DECODE) and variants of genome-wide significant in the sVEGFR2 pQTL of UKB-PPP study (Supplementary table 2).

### Selection of genetic instruments

All above mentioned datasets of sVEGFR2 pQTL (namely Internal, AGES, DECODE, Meta-analysis, and UKB-PPP study) were used to select genetic instruments for sVEGFR2. Genetic variants associated with sVEGFR2 with genome-wide significance and located ± 100 kilobase (kb) of *VEGFR2* gene (chromosome 4, GRCh38 position 55078481-55125595) were selected. These variants were then pruned to remove SNPs in high linkage disequilibrium (R^2^ < 0.1) within a 100kb window based on the 1000 Genomics European reference panel. To select variants associated with MP, a GWAS-correlated P-value < 5 × 10^−8^ and linkage disequilibrium (LD) R^2^ < 0.001 within a 10,000kb window based on the European 1000 Genome Project reference panel. Variants with F-statistic < 10 and minor allele frequency (MAF) < 0.01 were excluded. F-statistic > 10 indicates robust instrument strength, calculated by F = R^2^ (N - K - 1)/K(1 - R^2^), where R^2^ is the proportion of variance in the exposure explained by genetic variants, K is the number of instruments, and N is the sample size. R^2^ estimates the proportion of variance in the phenotype explained by the genetic variant, R^2^ = 2 × (Beta/SD)^2^ × (MAF) × (1 − MAF), where Beta represents the per-allele effect size of the association between each variant and the phenotype, and SD denotes the standard deviation (33). Detailed information on the genetic variants was provided in Supplementary table 3.

### *cis*-MR analysis

We performed MR analyses following the STROBE-MR guidelines (34). MR study should be conducted based on three core principles: (1) relevance, the genetic instruments should be significantly associated with the exposure; (2) independence, the genetic instruments should not be associated with any potential confounder and (3) exclusion restriction, the genetic instruments should not directly affect the outcome except via the way of exposure (18). Genetic correlations between exposure and outcome for each variant were harmonized by aligning effects alleles, with exclusion of palindromic variants. Prior to each MR analysis, MR Pleiotropy RESidual Sum and Outlier (MR-PRESSO) was applied to identify and remove potential outliers (35). The causal effects were assessed mainly using inverse variance-weighted (IVW) method of MR (36). This method assumes that outcomes are influenced only by the exposure, setting the intercept at zero (36). We also performed MR Egger, which allows sensitivity tests to rule out potential bias induced by horizontal pleiotropy using the inclusion of the intercept in the regression analysis, and P < 0.05 was considered significant for the presence of horizontal pleiotropy (37). MR-PRESSO global test was used to assess the presence of horizontal pleiotropy and P value < 0.05 was considered significant for the presence of horizontal pleiotropy (35). The Cochran’s Q test was used to assess heterogeneity, and P < 0.05 was considered significant for the presence of heterogeneity (38). The leave-one-out plots were used to assess the influence of individual variants on observed associations (38). The MR Steiger directionality tests were used to assess whether there was reverse causality in our *cis*-MR study (39). Reverse MR with IVW method aimed to evaluate whether MP was causally associated with sVEGFR2 levels and was employed. No proxy variants were used across the MR analyses.

### Bayesian colocalization analysis

Bayesian colocalization was performed to investigate whether sVEGFR2 and MP shared genetic variants within ±100 kb around the *VEGFR2* gene. The “coloc” package (version 5.2.3) in R (version 4.4.1) was utilized for these analyses (40), with visualization facilitated by the “locuscomparer” package (version 1.0.0) (41). The posterior probability of hypothesis 4 (PPH4) > 0.8 indicated strong evidence of colocalization.

### Statistical analysis

The IVW method was used to assess the causal effect. MR results were reported as odds ratios (OR) with 95% confidence interval (CI) per one-SD increase in log-transformed unit of sVEGFR2 levels. All MR analyses were two-sided and performed using package ‘TwoSampleMR’ (version 0.6.4) in R (version 4.4.1). P < 0.05 was considered statistically significant.

## Results

### Discovery of novel sVEGFR2-associated variants through genome-wide association meta-analysis

The overall study design was shown in Figure 1. Our meta-analysis on sVEGFR2 pQTL of Interval, AGES, and DECODE studies identified 30 independent variants with genome-wide significance. We mapped their corresponding genes and searched their CADD scores using Open Targets (31). Among these, six variants were considered novel, as they located outside the ±500 kb window of known loci identified in the original studies (Interval, AGES, and DECODE). After further excluding variants with genome-wide significance found in the UKB-PPP study, which included 33822 European individuals with over 20 million variants and used Olink technology for plasma protein measurement (26), four novel variants remained. Specifically, these were rs9411335 (*MED27*, Mediator Complex Subunit 27), rs6992534 (*ERI1*, Exoribonuclease 1), rs11893824 (*LIMS2*, LIM Zinc Finger Domain Containing 2), and rs7678559 (*PCDH7*, Protocadherin 7) (Table 1 and Supplementary figure 1).

**Figure 1.**
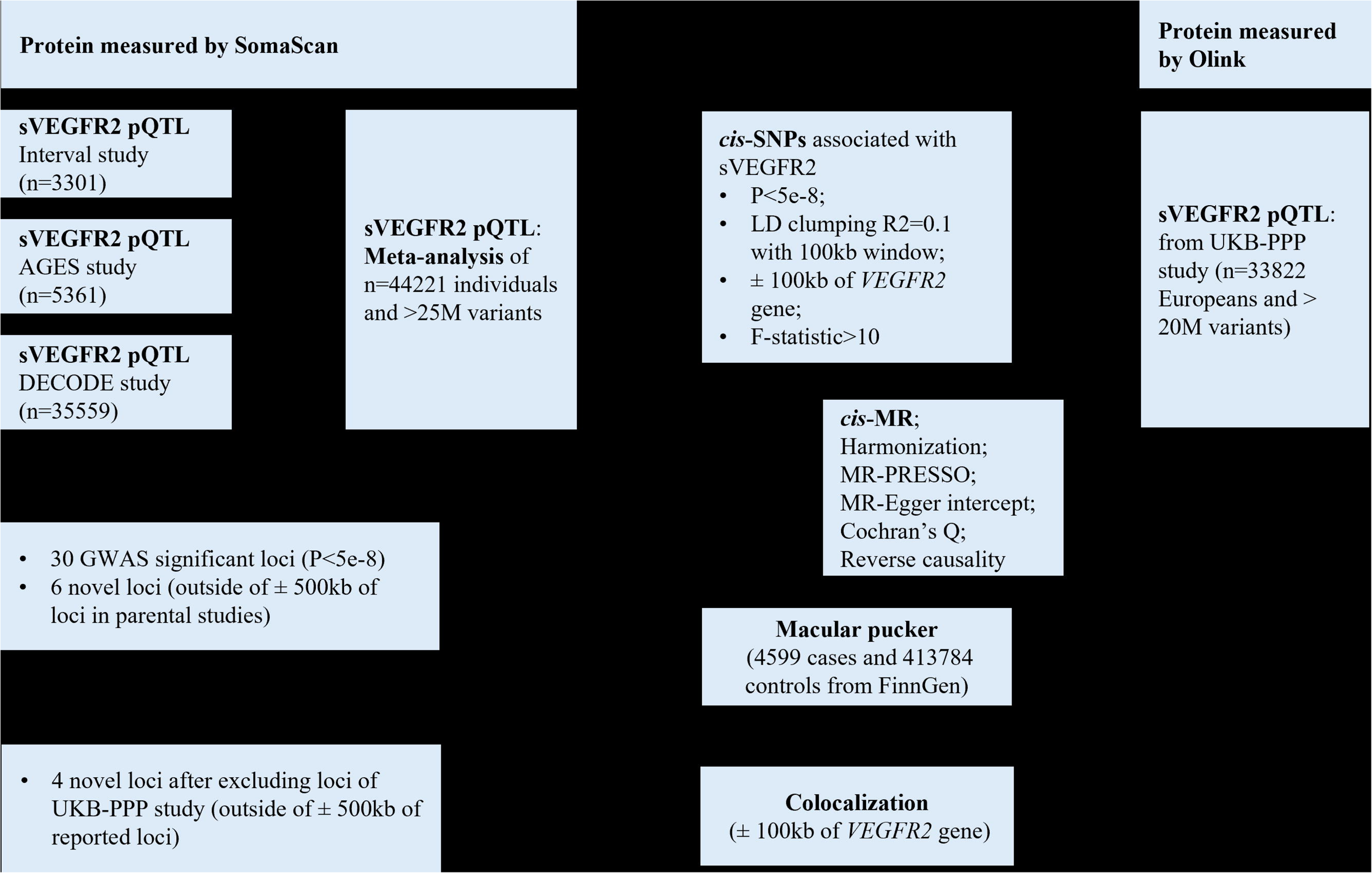
Study design and datasets. sVEGFR2, soluble vascular endothelial growth factor receptor 2; pQTL, protein quantitative trait loci; GWAS, genome-wide association studies; MR, mendelian randomization; MR-PRESSO, MR Pleiotropy RESidual Sum and Outlier; LD, linkage disequilibrium; SNP, single nucleotide polymorphism; Interval study, Sun BB. et al. 2018 Nature (ref. 23); AGES study, Gudjonsson A. et al. 2022 Nature Communications (ref. 24); DECODE study, Ferkingstad E. et al. 2021 Nature Genetics (ref. 25); UKB-PPP (UK Biobank Pharma Proteomics Project) study, Sun BB. et al. 2023 Nature (ref. 26).

**Table 1.**
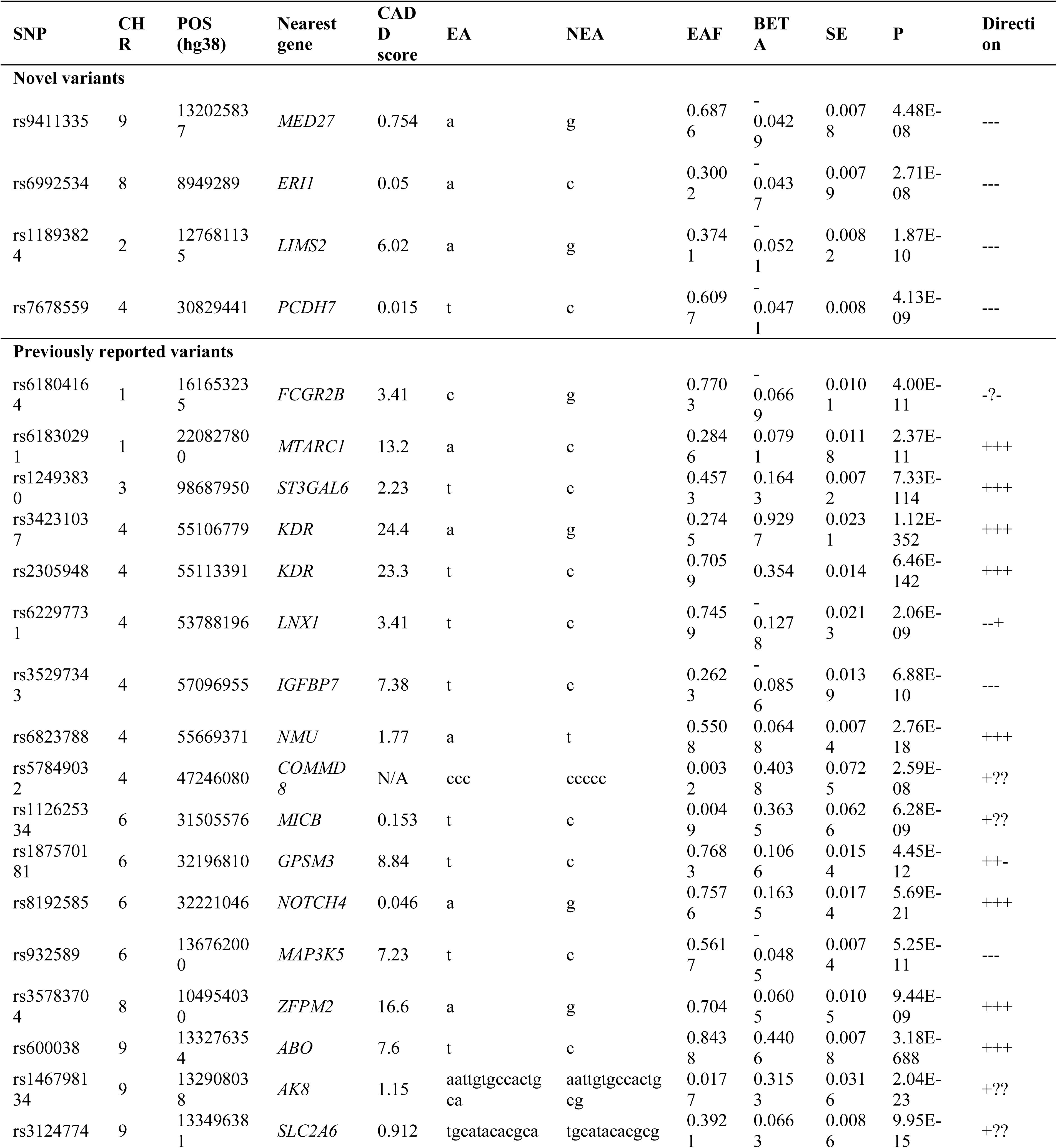
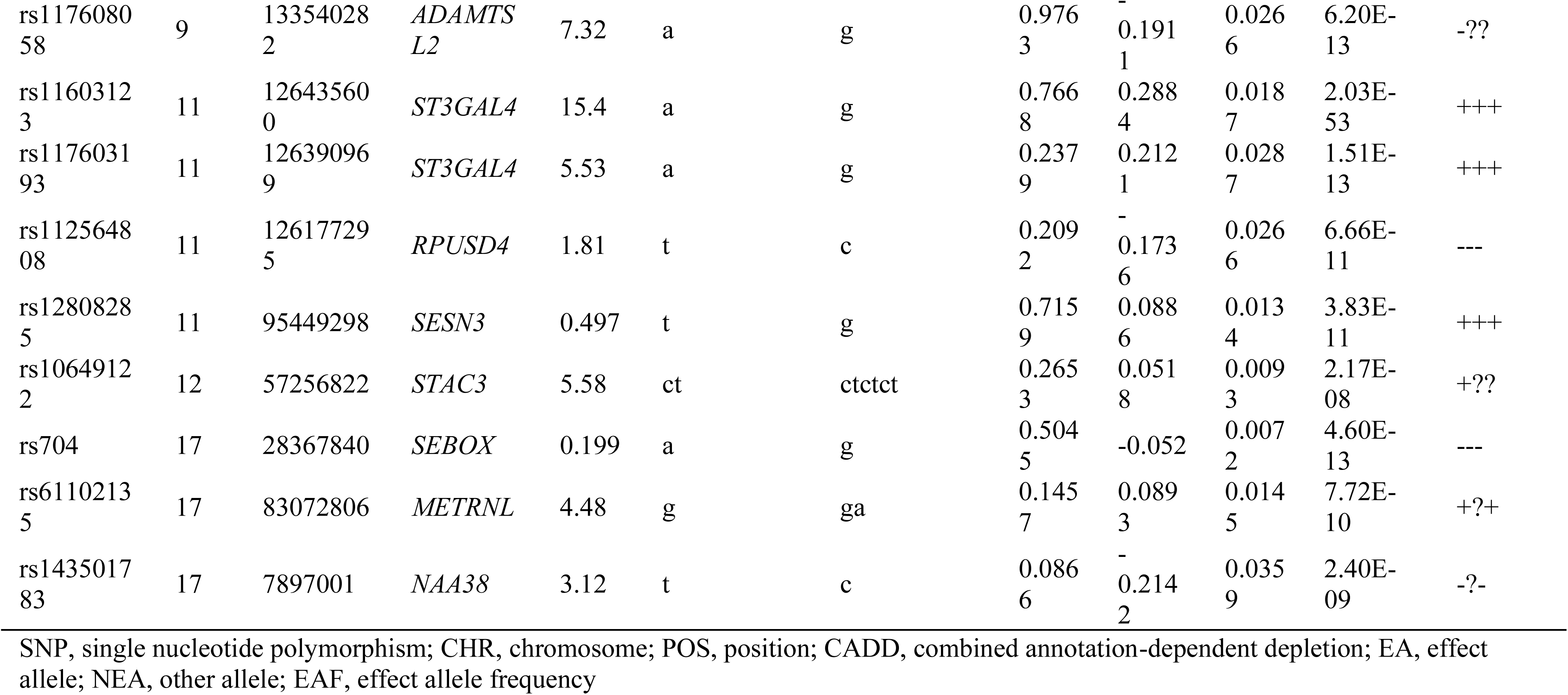
Variants reported for plasma levels of sVEGFR2 in the meta-analysis of Interval, AGES, and DECODE datasets.

### 3.2 *Cis*-MR and colocalization suggested sVEGFR2 as a protective factor against MP

*Cis*-MR analysis using IVW method indicated that genetically predicted higher levels of sVEGFR2 were associated with a reduced risk of MP (Figure 2). Specifically, for sVEGFR2 levels inferred from genetic variants in the meta-analysis, the odds ratio (OR) for MP was 0.86 (95% CI: 0.77-0.95, P = 4.6 × 10^−3^), while for sVEGFR2 levels inferred from the UKB-PPP study, the OR was 0.86 (95% CI: 0.78-0.96, P = 4.4 × 10^−3^). Additionally, sVEGFR2 levels predicted by genetic variants from the Interval and DECODE studies, but not the AGES study, also showed an inverse association with MP (Figure 2 and Supplementary figure 2).

**Figure 2.**
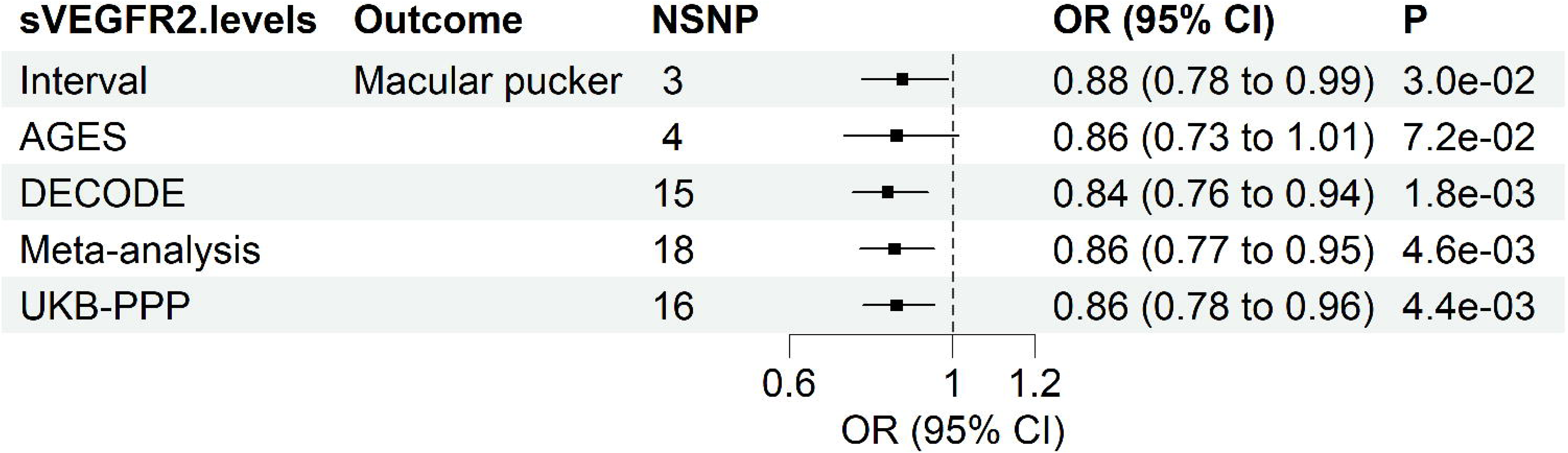
*Cis*-MR results of IVW method for the effects of sVEGFR2 levels on macular pucker. Results were reported as odds ratio (OR) with 95% confidence interval (CI) per one-standard deviation increase in sVEGFR2 levels; statistical tests were two-tailed; P < 0.05 was considered significant; IVW, Inverse variance weighted; NSNP, number of single nucleotide polymorphism; Meta-analysis, combined data of Interval, AGES, and DECODE studies.

To validate these findings, we conducted Bayesian colocalization analyses (Figure 3). The results revealed that the *VEGFR2* gene region was shared between sVEGFR2 pQTL and MP across all analyses. The PPH4 was as follows: 0.93 for the Interval study, 0.93 for the AGES study, 0.92 for the DECODE study, 0.92 for the meta-analysis, and 0.95 for the UKB-PPP study (Figure 3).

**Figure 3.**
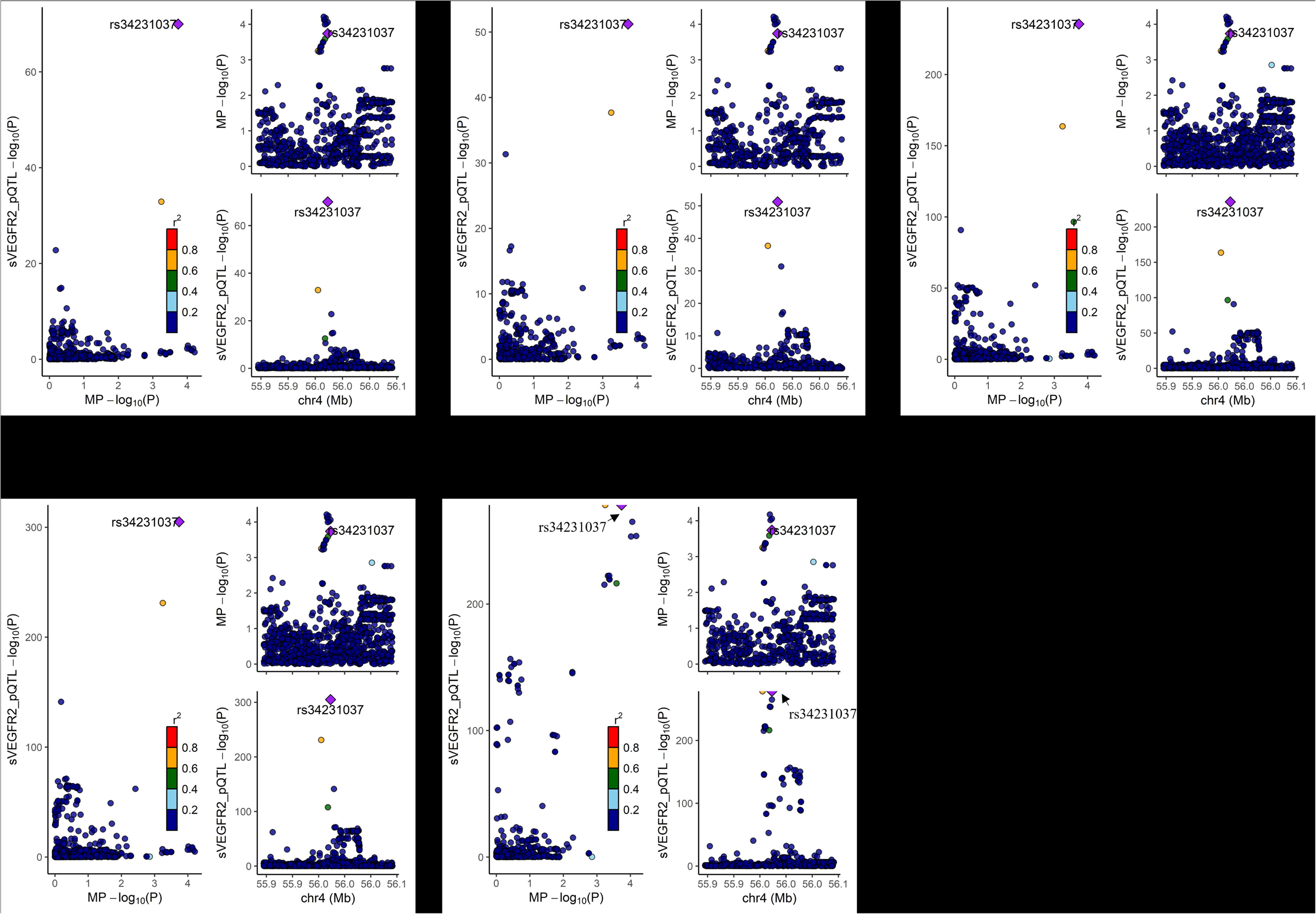
Colocalization of sVEGFR2 pQTL and macular pucker. Bayesian colocalization analysis for sVEGFR2 pQTL from Interval (A), AGES (B), DECODE (C), meta-analysis (D), and UKB-PPP (E) and macular pucker within ± 100 kb around the *VEGFR2* gene (chromosome 4, position 55,078,481-55,125,595). A posterior probability of hypothesis 4 (PPH4) > 0.8 indicated strong evidence of colocalization. In the regional plots, each dot represented a genetic variant, and the candidate causal variant was depicted as a purple diamond. The color of other variants indicated their linkage disequilibrium (r²).

### Tests for sensitivity and reverse causality

To ensure the robustness of our findings, we conducted a series of rigorous sensitivity analyses in our *cis*-MR study to examine the relationship between sVEGFR2 levels and MP risk. We employed MR-Egger intercept and MR-PRESSO global tests to assess horizontal pleiotropy. Both tests consistently showed no evidence of horizontal pleiotropy across all examined associations (Supplementary table 4). However, Cochran’s Q tests revealed significant heterogeneity between sVEGFR2 levels in the meta-analysis and the UKB-PPP study, and the outcome in our *cis*-MR analyses (Supplementary table 4). Despite this, the heterogeneity was deemed acceptable given that our primary *cis*-MR results were estimated using the IVW method. Additionally, leave-one-out plots suggested that no single genetic variant disproportionately influenced the overall results of our *cis*-MR study (Supplementary figure 3).

To address the potential for reverse causality, we performed MR Steiger directionality tests, which provided no evidence of reverse causation in the significant *cis*-MR results (Supplementary table 5). Further, we conducted reverse MR analyses, treating MP as the exposure and sVEGFR2 levels as the outcome. These analyses revealed that only the DECODE study and the meta-analysis contained genetically valid variants that also intersected with MP (Supplementary table 6). Importantly, no association between MP and sVEGFR2 levels was found, reinforcing the absence of reverse causality in our initial *cis*-MR findings (Supplementary table 6).

## Discussion

We presented a large-scale GWAS meta-analysis of SomaLogic values for plasma sVEGFR2, which enabled the discovery of four novel variants associated with sVEGFR2. This meta-analysis, along with an independent dataset for sVEGFR2 pQTL (measured by Olink) in the UKB-PPP study, further enabled MR analysis sourcing *cis*-variants within and around the *VEGFR2* gene to explore potential associations between sVEGFR2 and MP. Furthermore, the *cis*-MR results were supported by Bayesian colocalization analyses, showing the presence of shared genetic variants between sVEGFR2 and MP. To the best of our knowledge, our study provided the first human causal evidence that higher sVEGFR2 levels were associated with a reduced risk of MP. Therefore, these analyses proposed sVEGFR2 as a promising target for preventing or treating MP.

VEGFR2, a receptor primarily known for its role in endothelial cells, is also expressed in the retinal pigment epithelium (RPE) (7–9). RPE is a crucial monolayer located between the neuroretina and choroid and is implicated in the pathogenesis of MP (42). Dysregulation of VEGFR2 signaling in the RPE can lead to aberrant endoplasmic reticulum stress, oxidative stress, and inflammatory responses (43), all of which could be critical factors in MP development. Abnormal blood vessel formation is also observed in vascular MP, highlighting the significance of VEGFR2’s functions beyond its canonical roles in endothelial cells. VEGFR2’s traditional functions in endothelial cells include regulating blood vessel formation and permeability (10). These functions may also influence MP pathogenesis in the retina. The protective role of sVEGFR2 in MP is likely multifaceted. As a decoy receptor, sVEGFR2 binds VEGF-A, competing with membrane-bound VEGFR2 (mVEGFR2) (12). This interaction can modulate downstream processes such as angiogenesis, lymphangiogenesis, inflammation, and fibrotic responses in the macula (11), potentially preventing or alleviating MP progression. By sequestering VEGF-A, sVEGFR2 can also mitigate VEGF-A/mVEGFR2-mediated vascular permeability, contributing to retinal vasculature stabilization and potentially reducing MP pathogenesis (10). Further research is essential to fully elucidate sVEGFR2’s protective mechanisms in MP and to explore its potential therapeutic applications.

In addition, the meta-analysis identified four novel genetic variants associated with sVEGFR2: rs9411335 (MED27, Mediator Complex Subunit 27), rs6992534 (ERI1, Exoribonuclease 1), rs11893824 (LIMS2, LIM Zinc Finger Domain Containing 2), and rs7678559 (PCDH7, Protocadherin 7). Searches in the GWAS catalog did not reveal any studies linking these variants to sVEGFR2 or mVEGFR2. Moreover, the relationship between mVEGFR2, which is expressed on the cell surface, and the levels of sVEGFR2 secreted into circulation remains poorly understood. Further investigation of these variants could reveal important regulation mechanisms of VEGFR2 signaling and related diseases.

It has been observed that MP can be linked to various macular conditions (2). To explore whether sVEGFR2 might influence MP through its association with these conditions, we examined whether sVEGFR2 was related to several macular disorders. Interestingly, our analysis, using genetic variants identified from meta-analyses or the UKB-PPP study to predict sVEGFR2 levels, found no significant association with a range of macular conditions, including macular hole, retinal detachments and breaks, diabetic retinopathy, retinal vascular disorders, and retinal vascular occlusions (Supplementary figure 4). These findings suggested that the association between sVEGFR2 and MP may be specific to MP itself, rather than being influenced by a broader relationship with these other macular conditions.

Our study had several major strengths. We utilized a GWAS meta-analysis to generate a large-scale sVEGFR2 pQTL and further validated these findings with an independent large-scale sVEGFR2 pQTL from the UKB-PPP study. These studies represent plasma protein measurements obtained via SomaScan and Olink, respectively. The integration of these two datasets not only maximized the inclusion of genetic variants associated with sVEGFR2 but also significantly improved the robustness and reliability of our findings by complementing each other. Additionally, genetic variants were selected from regions ± 100kb of *VEGFR2* gene. These variants were likely to have a significant impact on protein expression compared to their impact on other traits. Therefore, *cis*-MR was less likely to violate the assumption of ‘no horizontal pleiotropy’. Besides, utilizing *cis*-MR for protein risk factors could enhance the reliability of causal inferences by aligning with Crick’s central dogma. This principle could support a sequential flow from gene to protein to disease, which could be more plausible than a gene to disease to protein pathway, particularly in studies using population-based samples. Therefore, *cis*-MR could effectively minimize the likelihood of reverse causality. Furthermore, Bayesian colocalization analysis is a powerful method used to determine whether two different traits or genomic signals are influenced by the same causal genetic variants. Integrating *cis*-MR with colocalization could synergistically harness their respective strengths, reinforcing the causal relationship between sVEGFR2 and MP.

To appreciably understand the results, several limitations in the study should be discussed. While we identified genetic variants significantly associated with sVEGFR2 levels, they explained only a small proportion of the total variance and should not be considered as exact proxies of the exposure. Moreover, MR effect estimates assume lifelong exposure to altered protein levels (e.g., sVEGFR2), which could potentially be diverging from observational associations and therapeutic interventions. Besides, sVEGFR2 levels measured in plasma from the published GWAS pQTL analysis may not accurately represent their concentrations or biological functions in retinal tissues, where they could potentially play a pivotal role in MP pathogenesis. This indicated the need for cautious interpretation of our findings. Our study primarily highlighted the causally protective effects of sVEGFR2 on MP, yet the underlying biological mechanisms require further investigation. Lastly, our study included only individuals of European descent, which could limit the generalizability to other populations.

## Conclusion

In conclusion, our study employed GWAS meta-analysis of sVEGFR2 pQTL and utilized *cis*-MR and colocalization analyses to provide robust genetic evidence supporting a protective effect of sVEGFR2 against MP. These findings suggested sVEGFR2 as a promising drug target for MP and warranted future basic and translational research in this area.

## Data Availability

All GWAS data are publicly available.

https://www.decode.com/

https://www.finngen.fi/en

https://gwas.mrcieu.ac.uk/

## List of abbreviations

MP: macular pucker
sVEGFR2: soluble vascular endothelial growth factor receptor 2
mVEGFR2: membrane-bound VEGFR2
GWAS: genome-wide association studies
pQTL: protein quantitative trait loci
MR: mendelian randomization
IVW: inverse variance weighted
MED27: mediator complex subunit 27
ERI1: exoribonuclease 1
LIMS2: LIM zinc finger domain containing 2
PCDH7: protocadherin 7
UKB-PPP: UK Biobank Pharma Proteomics Project
CADD: combined annotation-dependent depletion

## Ethical statement

In this study, we utilized publicly available GWAS data and ethical approvals were obtained in all original studies.

## Author contributions

Study design: J.G., Q.G.; data acquisition and analysis: J.G., Q.G.; figures, tables and original draft: J.G.; writing, reviewing and editing: J.G., Q.G.

## Data sharing statement

All GWAS data are publicly available and described in the data source section and supplementary table 1.

## Supplementary materials

Supplementary materials associated with this article can be found in the online version, at xxx.

---

**Supplementary figure 1.**
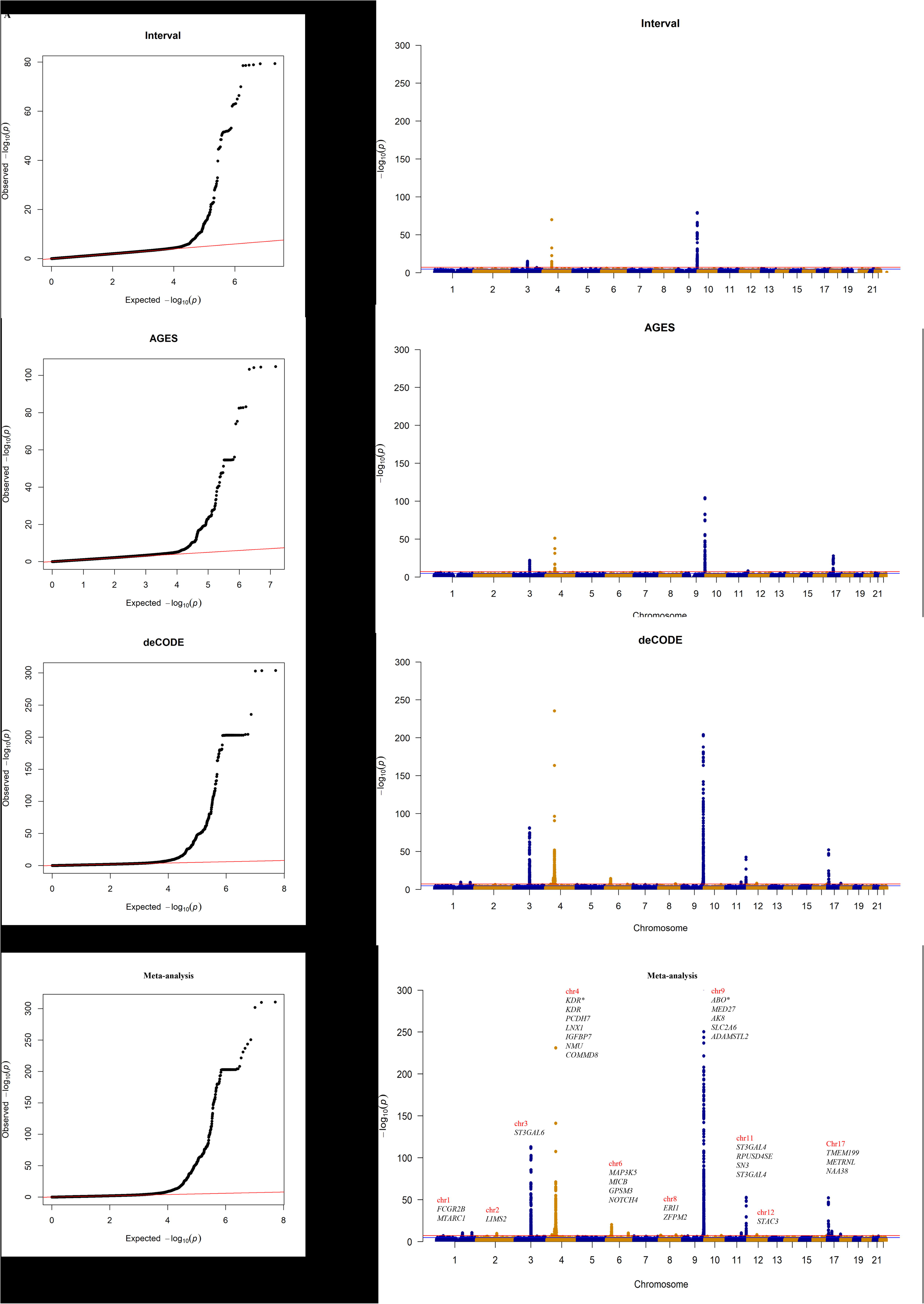
Q-Q plots and Manhattan plots of the sVEGFR2 pQTL. Mapped genes for GWAS-wide significant variants (P<5 × 10^−8^) in the Manhattan plot for the meta-analysis of the three independent studies were shown.

**Supplementary figure 2.**
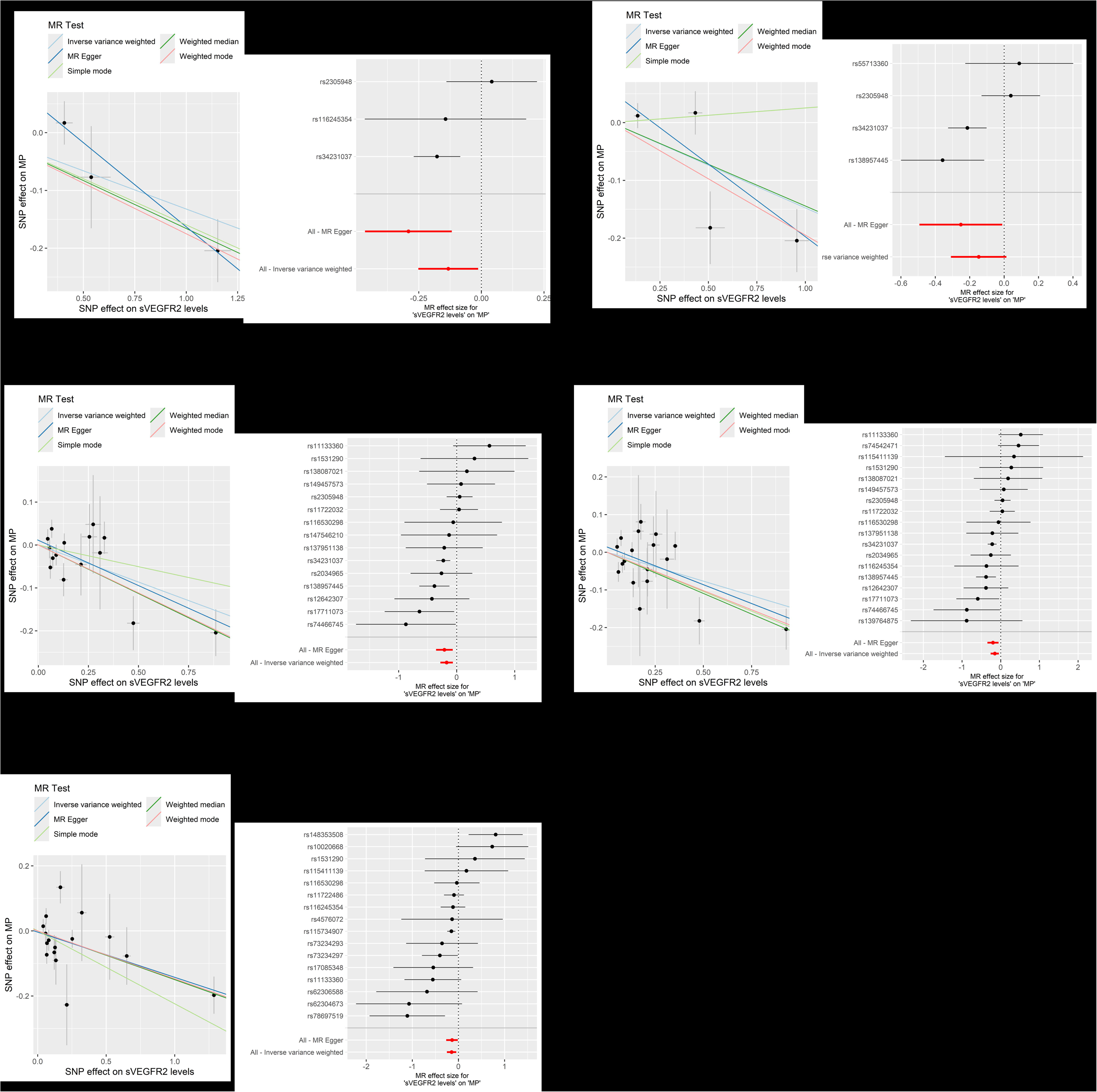
Scatter plots and forest plots in *cis*-MR analysis. In scatter plots, the x-axis shows the effect of each SNP on sVEGFR2 levels. The y-axis shows the effect of each SNP on macular pucker (MP). The regression lines for inverse variance weighted (IVW), weighted median, MR-egger, simple mode, weighted mode method are shown. In forest plots, the x-axis shows the *cis*-MR effect size for sVEGFR2 levels on macular pucker. The y-axis shows the analysis for each of the SNPs and for the SNPs in total using the MR-egger and IVW methods. The data are presented as raw β values with corresponding 95% confidence intervals. (A) Interval sVEGFR2 on MP; (B) AGES sVEGFR2 on MP; (C) DECODE sVEGFR2 on MP; (D) Meta-analysis sVEGFR2 on MP; (E) UKB-PPP sVEGFR2 on MP.

**Supplementary figure 3.**
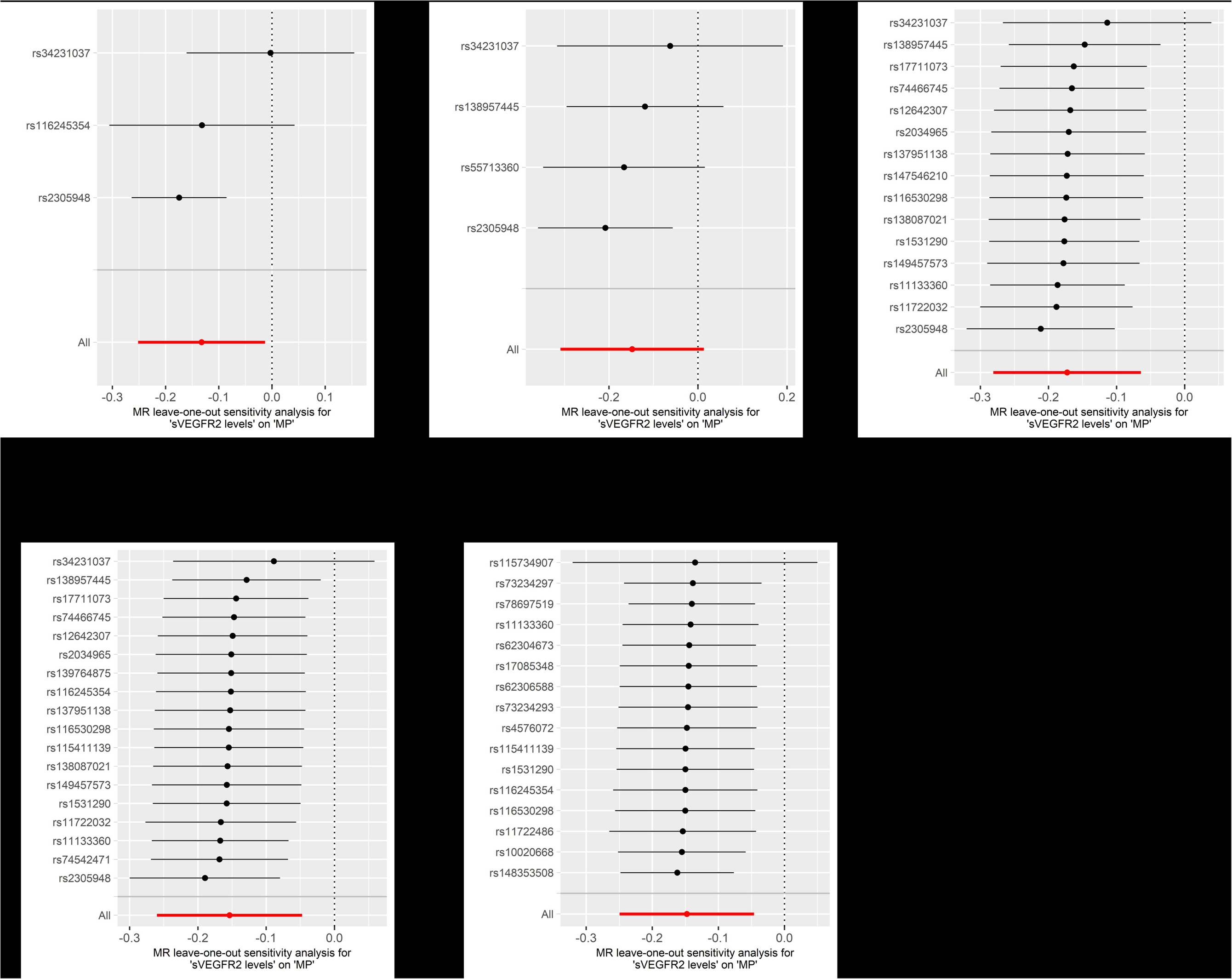
Leave-one-out plots of cis-MR analysis for the effect of sVEGFR2 levels on macular pucker. The x-axis shows the *cis*-MR leave-one-out sensitivity analysis for the effect of sVEGFR2 levels on macular pucker. The y-axis shows the analysis for leave-one-out of SNPs and the effect of the total SNPs on macular pucker. The data are presented as raw β values with corresponding 95% confidence intervals.

**Supplementary figure 4.**
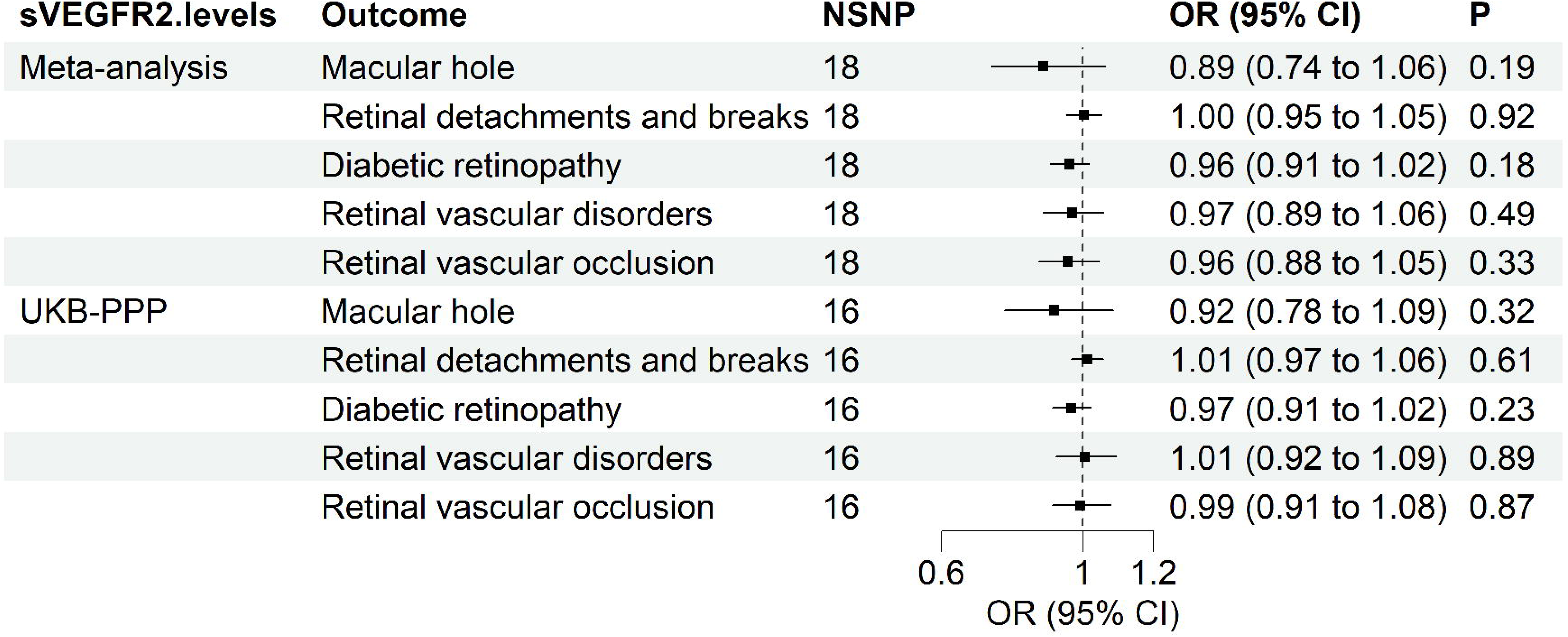
*Cis*-MR analysis using IVW method for the effects of sVEGFR2 levels on other eye diseases. Results were reported as odds ratio (OR) with 95% confidence interval (CI) per one-standard deviation increase in sVEGFR2 levels; statistical tests were two-tailed; P < 0.05 was considered significant; IVW, Inverse variance weighted; NSNP, number of single nucleotide polymorphism; Meta-analysis, combined data of Interval, AGES, and DECODE studies.

